# Circulating metabolites and physical performance are predictors of overall survival in metastatic lung cancer patients

**DOI:** 10.1101/2023.08.23.23294489

**Authors:** Willian das Neves, Christiano R. R. Alves, Gabriela dos Santos, Maria J. N. Alves, Amy Deik, Kerry Pierce, Courtney Dennis, Lily Buckley, Clary B. Clish, Kathryn J. Swoboda, Patricia C. Brum, Gilberto de Castro

**Affiliations:** Instituto do Cancer do Estado de Sao Paulo ICESP, Hospital das Clínicas HCFMUSP, Faculdade de Medicina, Universidade de Sao Paulo, Sao Paulo, Brazil; Center for Genomic Medicine, Massachusetts General Hospital, Boston, MA; Department of Neurology, Harvard Medical School, Massachusetts General Hospital, Boston, MA; School of Physical Education and Sport, University of São Paulo, São Paulo, Brazil; Heart Institute, School of Medicine, University of Sao Paulo, Sao Paulo, Brazil; Metabolomics Platform, Broad Institute of MIT and Harvard, Cambridge, MA; Current affiliation: Department of Physiology and Biophysics, Institute of Biomedical Science, University of Sao Paulo, Sao Paulo, Brazil

**Author notes:** **Corresponding author** Gilberto de Castro Junior, MD, PhD and Willian das Neves, PhD Faculdade de Medicina da Universidade de São Paulo, Medical Oncology - Instituto do Câncer do Estado de São Paulo, Av. Dr. Arnaldo 251, 5th floor, ZIP code: 01246-000, São Paulo-SP, Brazil. and.

**Keywords:** Non-small cell lung cancer, metabolomics, physical performance, cancer cachexia, skeletal muscle wasting

## Abstract

**Background:** Skeletal muscle atrophy and low physical performance are associated with disease progression and higher mortality rates in multiple pathological conditions. Here, we determined whether body composition and physical performance would predict mortality in metastatic non-small cell lung cancer (NSCLC) patients. In addition, we defined whether plasma samples from NSCLC patients would directly affect the homeostasis of skeletal muscle cells. **Methods:** The prospective cohort included 55 metastatic NSCLC patients and seven age-matched control subjects. We assessed clinical characteristics, body composition, cancer cachexia, and quality of life (QoL). We determined physical performance with a series of functional tests. We analyzed skeletal muscle and adipose tissue areas. Finally, we evaluated the overall survival rate, and additional blood samples were collected from a subcohort of eighteen patients for further studies in cell culture and metabolomic analysis. **Results:** We found that physical performance, not body composition, was associated with overall survival in this cohort. Moreover, incubation with plasma derived from NSCLC patients with low physical performance impaired the metabolism and proliferation of primary human myotubes. Unbiased metabolomics revealed several metabolites differentially expressed in the plasma of NSCLC patients with low physical performance compared to healthy control subjects, with serine and N2,N2-dimethylguanosine (M22G) being the most reduced and increased metabolites, respectively. **Conclusion:** These novel findings confirm physical performance as a significant predictor of overall survival in metastatic NSCLC patients and provide insights into cancer-induced circulating factors that can directly affect skeletal muscle homeostasis and prognosis.

## Introduction

Approximately 1.8 million people die every year due to lung cancer, making this disease the leading cause of cancer-related death worldwide ^1^. The most common histological type of lung cancer is non-small cell lung cancer (NSCLC), accounting for more than 80% of all cases. Approximately 70% of all NSCLC cases are diagnosed with advanced stages, high symptom burden, low physical performance, and a survival rate lower than 30% within the first year of diagnosis ^2–5^.

Immunotherapies and molecular-targeting therapies are not generalizable to all NSCLC patients and are still restricted in many countries. In these locations, platinum-based chemotherapy is the primary treatment option for metastatic NSCLC patients, with a response rate of only ∼19% ^6^. Notably, this chemotherapy regimen induces considerable toxicity, which can further compromise patients’ quality of life and physical performance ^3^. Moreover, approximately half of NSCLC patients have severe body weight loss (*i.e.,* cachexia) and fatigue at the diagnosis stage ^7^, limiting adequate treatment exposure. Depending on their performance status, these patients are unsuitable for chemotherapy ^8,9^, and there is no available pharmacological treatment for cancer cachexia ^10^. Therefore, there is a critical need to identify biomarkers to predict the early onset of symptoms and to apply better palliative care to preserve physical performance in NSCLC patients.

In the present study, we tested the hypothesis that skeletal muscle atrophy and low physical performance are associated with higher mortality rates in NSCLC patients. Moreover, we determined whether plasma samples from NSCLC patients would directly affect the homeostasis of skeletal muscle cells. We performed unbiased metabolomics in these samples to identify potential novel biomarkers to track NSCLC progression. This approach revealed factors differentially regulated in NSCLC patients with low physical performance, providing novel insights into cancer-induced circulating factors that can directly affect skeletal muscle homeostasis.

## Results

### Establishing a cohort of NSCLC patients for investigation

Our service (ICESP; Sao Paulo, Brazil) admitted 1079 lung cancer patients between March 2017 and September 2020. Eighty patients were invited, and 60 consented to participate in this study. We analyzed a final cohort of 55 NSCLC patients who fulfilled all eligibility criteria. The subjects’ flowchart is illustrated in **Figure 1**, and the baseline clinical characteristics of the 55 analyzed patients are presented in **Table 1**. The median longitudinal follow-up was 7.9 months, with a range from 0.3 to 36.4 months and an overall survival rate of 29.9% in one year.

**Figure 1.**
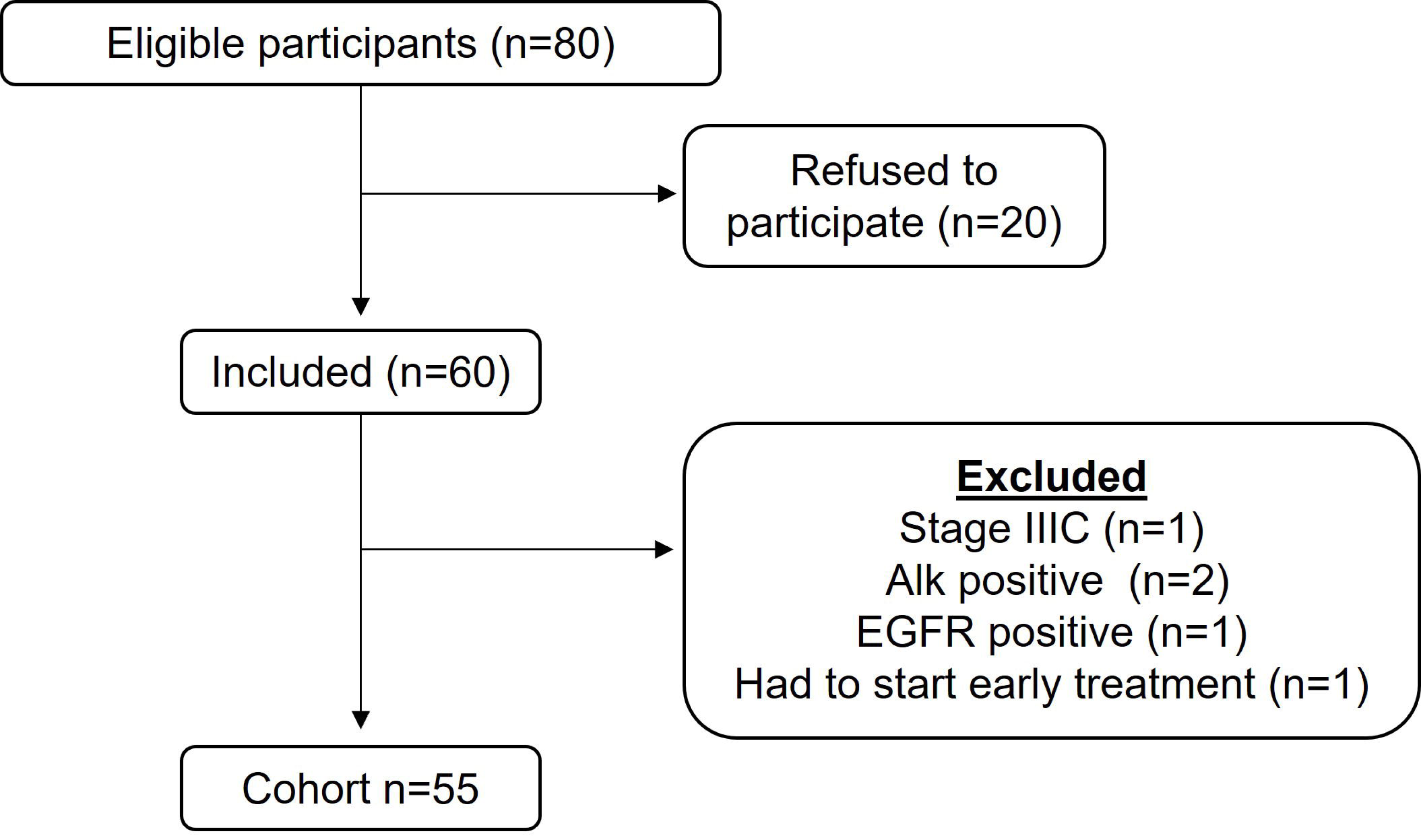
Flowchart detailing subject selection. NSCLC: non-small cell lung cancer.

**Table 1:** Patients Characteristics.

| Characteristics | N = 55 | % |
| --- | --- | --- |
| Age |  |  |
| Average (SD) | 65.5 (9.1) |  |
| Sex |  |  |
| Male | 48 | 87.3 |
| Female | 7 | 12.7 |
| Histology |  |  |
| Adenocarcinoma | 32 | 58.2 |
| SCC | 21 | 38.2 |
| NSCC | 2 | 3.6 |
| Cachexia |  |  |
| Yes | 38 | 69.1 |
| No | 16 | 29.1 |
| Unknow | 1 | 1.8 |
| Smoking Status |  |  |
| < 25 pack-years | 7 | 12.7 |
| 25-50 pack-years | 21 | 38.2 |
| 50-75 pack-years | 10 | 18.2 |
| > 75 pack-years | 10 | 18.2 |
| Smokers unknow amount | 7 | 12.7 |
| Outcomes |  |  |
| Dead | 41 |  |
| Overall survival in one year |  | 29.9 |
| Average of follow up in months | 7.9 |  |

### Physical performance, but not body composition, is a predictor of mortality in NSCLC patients

Since the Eastern Cooperative Oncology Group Performance Status (ECOG-PS) scale is a critical measurement in clinical practice to determine NSCLC patients’ physical ability ^11^, we first determined whether the ECOG-PS score was associated with overall survival in this cohort. We compared the survival rate across subjects with ECOG-PS status scales of 0, 1, and 2. As expected, better ECOG-PS scores were associated with higher survival rates (**Figure S1A**). Moreover, since cancer cachexia has been reported as a mortality predictor in other advanced cancers ^7,12^, we compared the survival rate between patients classified as cachectic or noncachectic at baseline. This comparison indicates a tendency for a higher survival rate in noncachectic patients than in cachectic patients, although it did not reach a statistically significant difference between groups (**Figure S1B**). Based on these initial findings, we performed similar analyses involving a series of actual body composition and physical performance measurements. We also analyzed the prognostic impact of blood cell counts on overall survival (**Supplementary Note** and **Figure S2**).

For each composition and physical performance assessment, we ranked the respective outcome into terciles (high, intermediate, and low) and compared the probability of survival across patients into each tercile. All body composition measurements were also compared across terciles, including the area of the psoas muscle, skeletal muscle index, total adipose tissue area, and intramuscular, subcutaneous, and visceral adipose tissue area (**Figure 2A-D** and **Table S1**). Physical performance measurements were significantly different across terciles, including scores in the timed up and go test, sit-to-stand test, six-minute walking test, and maximal oxygen uptake (**Figure 2E-H**). Better functional performance in these physical tests was associated with a higher survival rate (**Figure 2E-H**). Additional analysis related to physical performance included scores from different questionnaires, including variables related to fatigue, physical decline, and physical health (**Figure 2I-L**). These data indicate that body composition is not associated with survival rate in NSCLC patients, but physical performance predicts early mortality in these patients.

**Figure 2.**
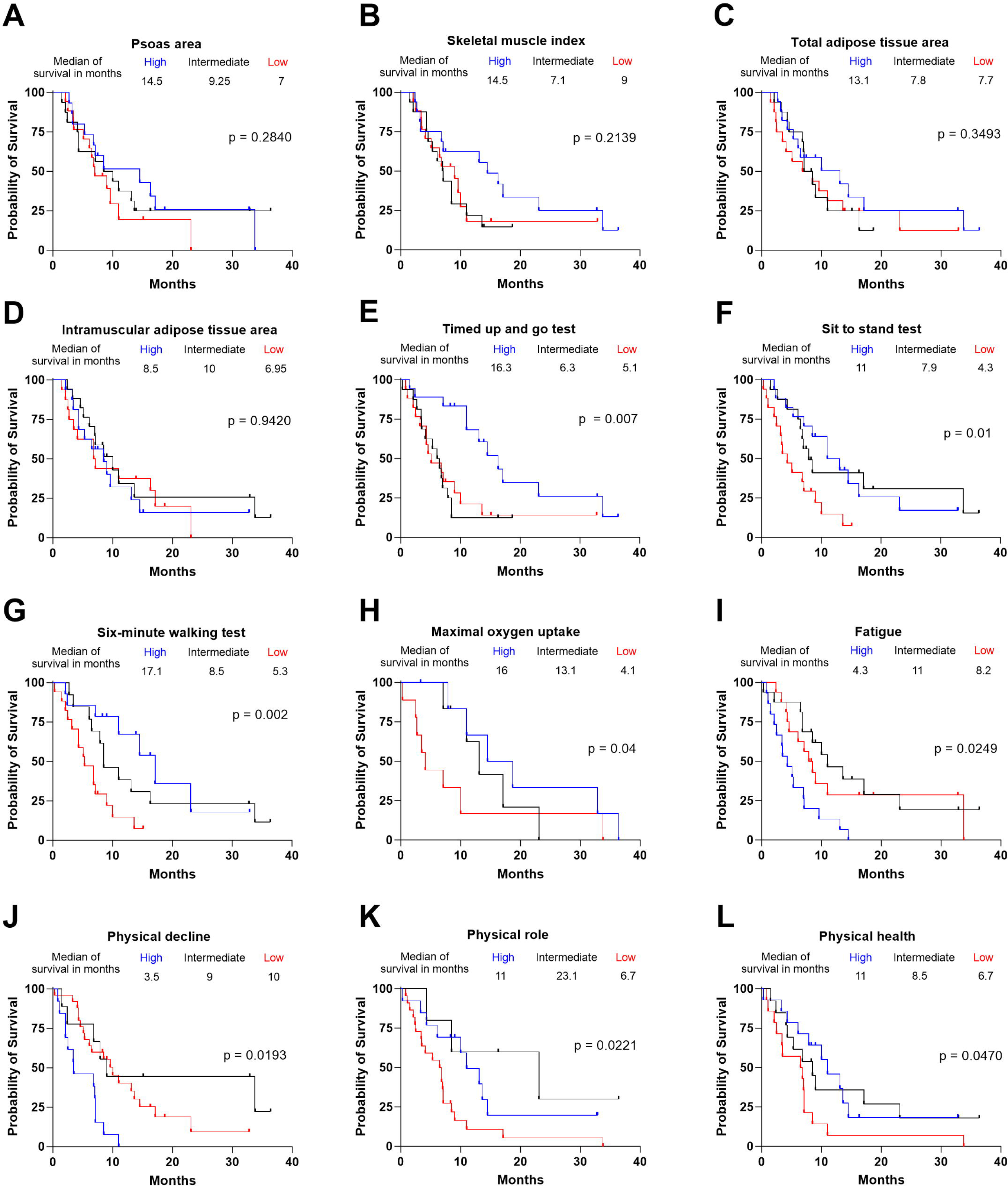
Associations between body composition, physical performance, and probability of survival in non-small cell lung cancer (NSCLC) patients. Each measurement was divided into tertiles (high, intermediate, and low), and tertiles’ probability of survival was compared. Body composition analysis included (**A**) the area of the psoas muscle, (**B**) skeletal muscle index, (**C**) total adipose tissue area, and (**D**) intramuscular adipose tissue area. Physical performance analysis included the (**E**) timed up and go test, (**F**) sit-to-stand test, (**G**) six-minute walking test, and (**H**) maximal oxygen uptake. Physical performance analysis based on questionaries included the (**I**) fatigue category from QLQ-C30, (**J**) physical decline from CAX24, (**K**) physical role from SF36, and (**L**) physical health from the SF36. Comparisons were performed using the log-rank test and presented as Kaplan Meier curves. n = 55 (except for maximal oxygen uptake, in which a subset of 24 subjects was analyzed).

### Severe metabolic impairment in muscle cells incubated with plasma from NSCLC patients

We have previously demonstrated that primary mouse myotubes exposed to conditioned media from cancer cells are metabolically stressed ^13^. To expand our knowledge on this topic, we used the present NSCLC cohort to determine the metabolic effects of circulating factors from NSCLC patients directly into healthy human myotubes. We collected samples from a subset (17/55) of NSCLC patients and included seven age-matched healthy control subjects to perform these studies (**Table S3**). Primary human myotubes were incubated for 16 hours with media containing 10% plasma collected from each subject. Some of these plasma samples affected the basal oxygen consumption rate (OCR) and extracellular acidification rate (ECAR) in healthy myotubes (**Figure S3**). Each readout was further analyzed after dividing subjects into terciles based on sit-to-stand performance to understand the association of these observed effects with physical performance. Remarkably, only plasma derived from patients with low physical performance affected basal OCR and ECAR in myotubes (**Figure 3A-B**). Although the OCR and ECAR data were normalized to the total protein content, we also observed that incubation with plasma derived from patients with low physical performance decreased the cell proliferation rate and total protein content in myotubes (**Figure 3C-D**). No difference was detected in OCR and ECAR between myotubes incubated with plasma from healthy control subjects and NSCLC patients with high physical performance (**Figure 3C-D**). Thus, incubation with plasma collected exclusively from NSCLC patients with low physical performance induced severe metabolic changes, reduced proliferation, and induced death and atrophy in myotubes.

**Figure 3.**
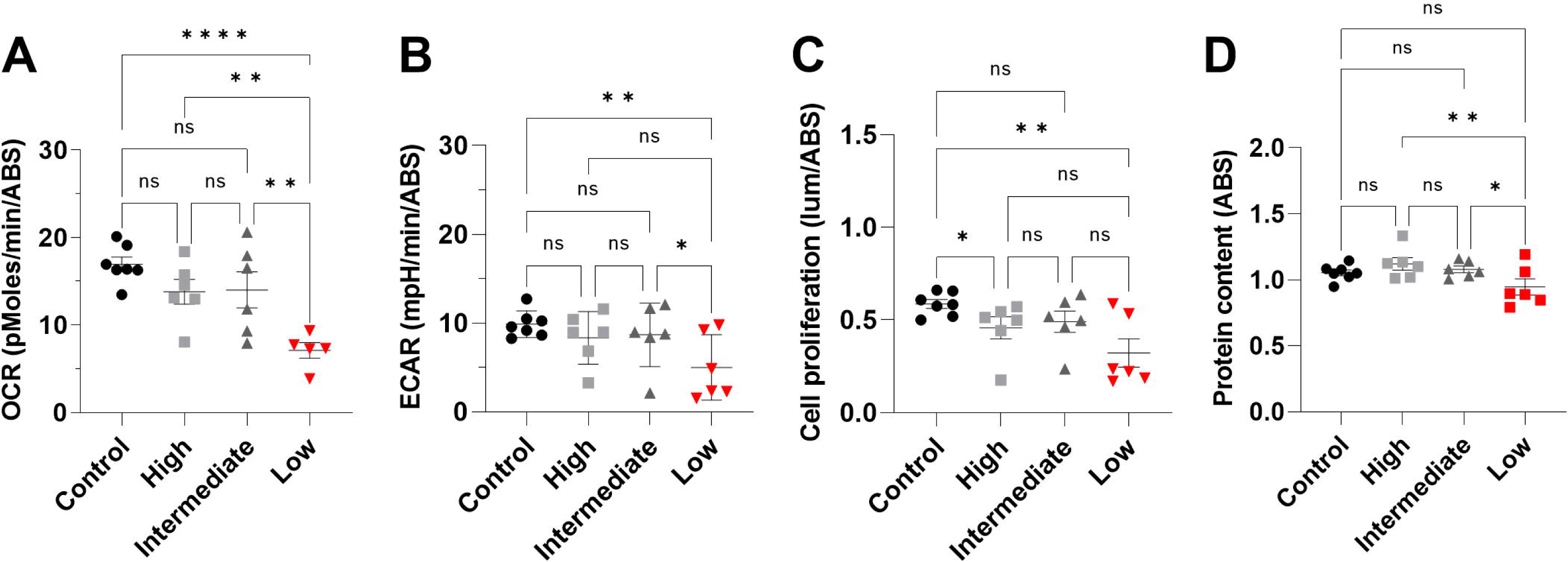
Metabolic changes in primary human myotubes treated with plasma from NSCLC patients. Differentiated primary human myotubes were incubated for 16 hours with media containing 10% plasma collected from each subject. Subjects were divided into tertiles based on physical performance from Figure 2F. (**A**) Oxygen consumption rate (OCR). (**B**) Extracellular acidification rate (ECAR). (**C**) Cell proliferation. (**D**) Protein content. Plots are derived from three independent experiments, and data are presented as mean ± s.e.m and individual values. *p < 0.05; **p < 0.01; ****p < 0.0001. n.s. not significant. n = 6-7.

For subsequent experiments, we selected plasma samples from 4 NSCLC patients with low physical performance who showed the most severe effects on myotubes (**Figure 3A-D**). In addition, we reduced the human plasma concentration in the media from 10% to 1.5% to avoid exacerbated cell death. Incubation with this reduced concentration did not affect myotube total protein content or nuclei counts (**Figure 4A-C**). However, we consistently observed reduced basal OCR and cell proliferation after incubation with plasma from NSCLC patients with low physical performance (**Figure 4D-E**). We also evaluated whether plasma from NSCLC patients with low physical performance could affect the activity of a direct repeat 4 (DR4) response element in C2C12 myoblasts, as we previously observed in murine models of lung cancer ^13^. As expected, there was reduced DR4 luciferase activity after treatment with plasma from these NSCLC patients (**Figure 4F**). Therefore, muscle cells incubated with a low plasma concentration from NSCLC patients with low physical performance have a consistent metabolic impairment.

**Figure 4.**
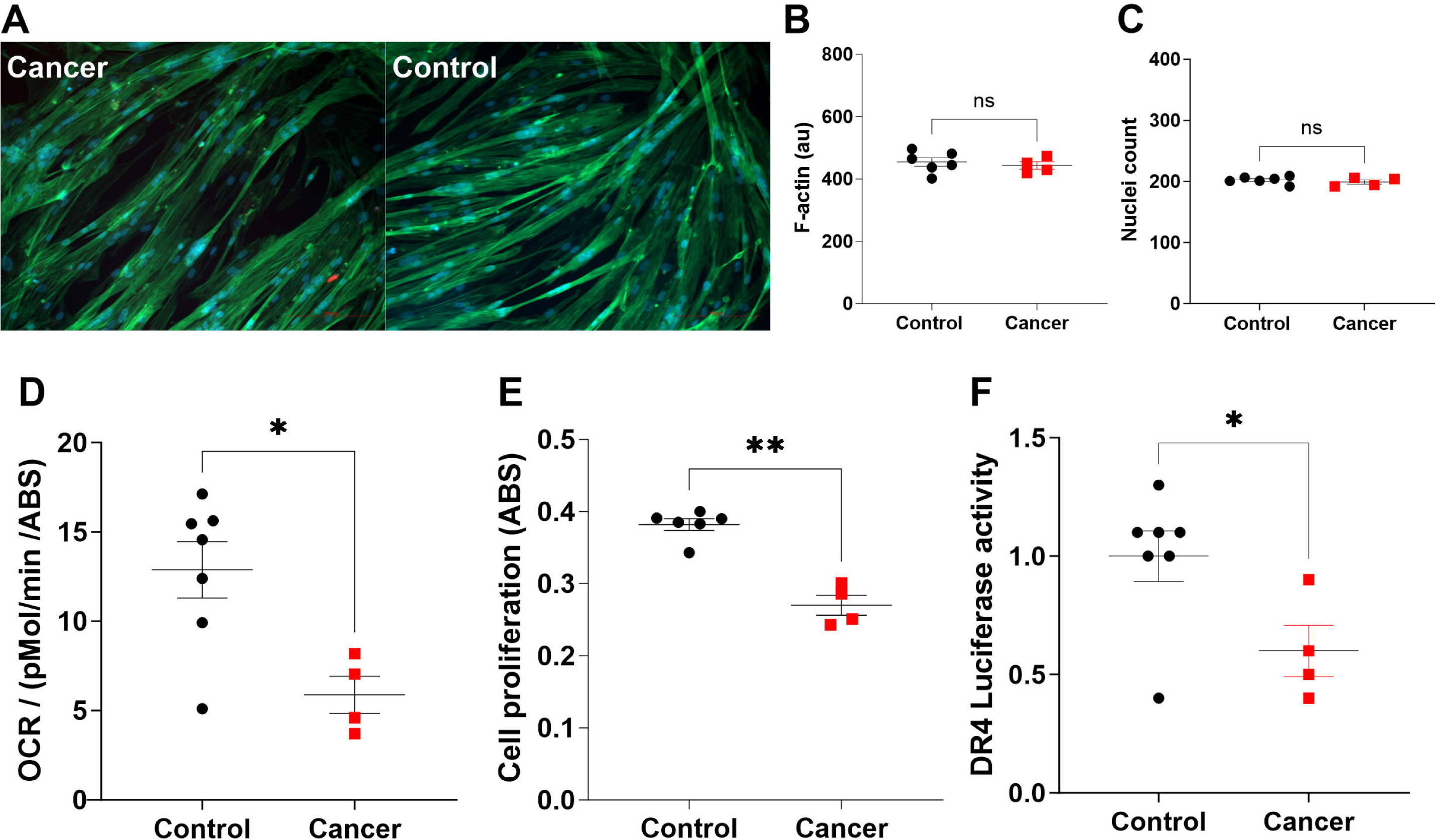
Metabolic changes and DR4 activity in muscle cells treated with a low percentage of plasma from NSCLC patients with low endurance capacity. Differentiated primary human myotubes (**A-E**) or C2C12 muscle cells (**F**) were incubated for 16 hours with media containing 2% plasma collected from each subject. A: Representative immunofluorescence for myotubes (20X) stained for F-actin and DAPI. B: Quantification of F-actin expression. C: Nuclei count. D: Oxygen consumption rate (OCR). E: Cell proliferation. F: Fold change of DR4 activity. Plots are derived from three independent experiments, and data are presented as mean ± s.e.m and individual values. *p < 0.05; **p < 0.01. n = 6-7.

### Changes in the plasma metabolite profile in NSCLC patients with low physical performance

Having established a set of plasma samples that directly affect myotube metabolism, we applied metabolomic screening to determine the plasma metabolites differentially expressed in these samples compared to healthy control subjects. We identified a total of 932 metabolites in all analyzed samples that were further divided into four categories: a) amino acids/acylcarnitines, b) free fatty acids, c) TCA cycle and glycolysis, and d) lipids (**Figure 5A**). By setting a cutoff of FDR at 10%, differentially expressed metabolites in NSCLC patients included 31 amino acids/acylcarnitines, one fatty acid, and three metabolites participating in the TCA cycle or glycolysis (**Figure 5A-B** and **Figure S3**). Of note, 13 metabolites in this list were sphingolipids and glycerophosphoethanolamines (**Figure S4**) and were all upregulated. Remarkably, the most significantly upregulated metabolite in NSCLC patients was N2,N2 dimethylguanosine (M22G), while the most downregulated metabolite was serine (**Figure 5B**). As expected, these two metabolites had the strongest inverse correlation (r = −0.99) across all differentially expressed metabolites in this list (**Figure 5C**).

**Figure 5.**
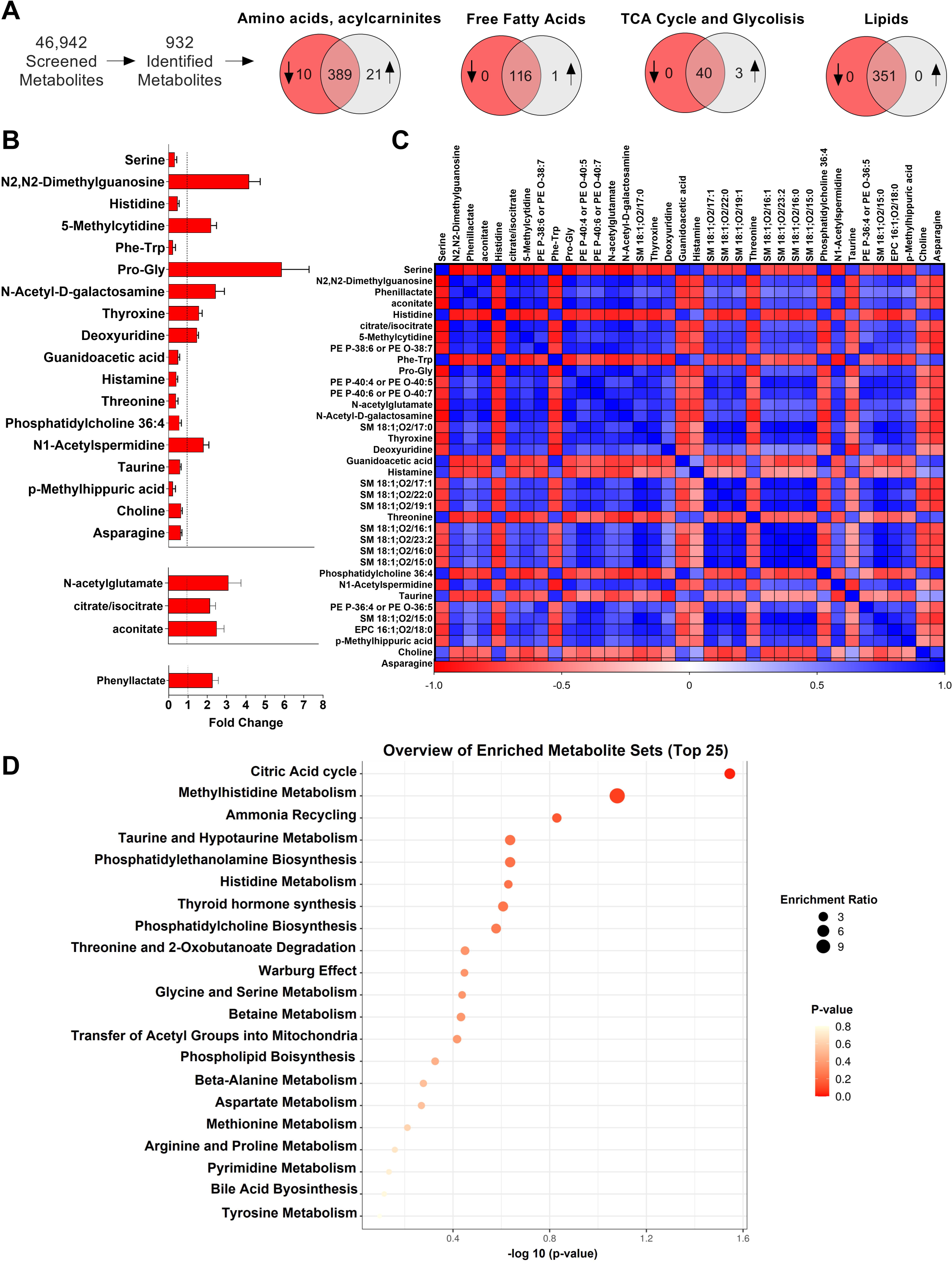
Plasma metabolomic profile in NSCLC patients with low endurance capacity. Metabolomic profiling was performed, and 932 identified metabolites were compared between NSCLC patients with low endurance capacity and control subjects. (**A**) The total number of differentially expressed metabolites between groups (FDR < 10%) was divided into four categories. (**B**) List of differentially expressed metabolites. (**C**) Correlation between each differentially expressed metabolite. (**D**) Pathway analysis (SBPNB) using all differentially expressed metabolites. n = 6-7.

The Small Molecule Pathway Database (SMPDB) and Kyoto Encyclopedia of Genes and Genomes (KEGG) pathway analysis inputting the differentially expressed metabolites in NSCLC patients revealed that the TCA cycle was the most affected pathway (**Figure 5D** and **Figure S5**), which corroborates the overall metabolic changes observed when treating myotubes with these plasma samples. The second most enriched pathway in this analysis was methylhistidine metabolism (**Figure 5D**), a metabolite produced in skeletal muscles during the methylation of actin and myosin. In summary, this plasma metabolite profile provides novel insights into circulating metabolites in NSCLC patients that could directly affect muscle metabolism.

## Discussion

Studies using animal models have demonstrated that preserved skeletal muscle mass and improved muscle oxidative metabolism can induce resistance to cancer progression ^14,15^. However, animal models have apparent limitations, and clinical evidence in this context is limited. Therefore, we collected data from a prospective cohort of metastatic NSCLC patients to study the potential of body composition and physical performance as predictors of mortality in these patients. Our findings indicate that the functional performance in different physical tests is associated with overall survival in metastatic NSCLC patients. Here, we also detected some tendency for the negative impact of the presence of cachexia on the overall survival of these patients, in line with a previous retrospective study of 134 metastatic NSCLC patients receiving chemotherapy ^7^. However, we did not observe a clear association between several body composition measurements and overall survival. In summary, these findings indicate that physical performance better predicts overall survival than body composition in NSCLC patients.

We collected plasma samples from a subset of NSCLC patients to determine whether circulating factors in NSCLC patients would directly affect muscle cells. Interestingly, we found that plasma samples collected from NSCLC patients with low physical performance affected metabolism and proliferation in primary myotubes. These findings not only corroborate previous studies demonstrating that conditioned media from murine lung cancer cells can induce stress in myotubes ^13,16–18^ but also demonstrate that these effects are associated with the physical condition of each NSCLC patient. Interestingly, samples from NSCLC patients with higher physical performance failed to promote the same catabolic effects in myotubes. Moreover, our group has previously identified that NSCLC patients have reduced muscle expression of COPS2, a known regulator of DR4 response elements ^13^. Here, we demonstrate for the first time that incubation with low concentrations of plasma from NSCLC with low physical performance reduces DR4 activity in muscle cells by ∼40%. Therefore, muscle cells incubated with plasma from NSCLC patients with low physical performance have consistent metabolic impairment, including mechanisms that may involve DR4 activity, which requires further investigation.

Plasma or serum metabolomics has been helpful in identifying key regulators of both cancer progression and muscle dysfunctions ^19–22^. Because the complex crosstalk between NSCLC and skeletal muscle has not been uncovered, we explored potential metabolic factors differentially expressed in the plasma of NSCLC patients with low physical performance. Unbiased metabolomic studies of these samples revealed that M22G is the most upregulated metabolite, while serine is the most downregulated in NSCLC patients with low physical performance. M22G is a purine nucleoside classified as a secondary metabolite. While its role in the human body is not fully understood, there is evidence that this metabolite helps to stabilize tRNAs ^23^.

Interestingly, M22G is present in tumors from NSCLC patients ^24^, and increased circulating M22G was previously described as a metabolite associated with different causes of death, including cancer-related mortality ^25–27^. Serine is a nonessential amino acid used in the biosynthesis of proteins and is also necessary for the functionality of muscle cells^28,29^. Thus, we speculate that the decreased circulating serine in NSCLC patients with low physical performance is associated with the muscle demand for this amino acid during the highly catabolic condition of these patients. These data provide novel insights into cancer-induced circulating factors that can directly affect skeletal muscle metabolism and uncover M22G and serine as putative biomarkers to track NSCLC progression. Further studies are encouraged to characterize the expression of these two metabolites in NSCLC patients.

In summary, these novel data establish physical performance as a significant predictor of overall survival in NSCLC patients and provide insights into cancer-induced circulating factors that can directly affect skeletal muscle homeostasis. These physical functional tests or metabolic measurements must be validated as potentially valuable biomarkers in future clinical trials in NSCLC patients for better stratification. Finally, additional challenging studies are needed to investigate whether ameliorating physical performance would increase the overall survival rate in NSCLC patients as a nonpharmacological intervention.

## Methods

### Ethics, Subjects, and Study Design

This study was approved by the Human Research Ethics Committee at the University of São Paulo (CEP-FMUSP; protocol: 2.286.563) and registered in *ClinicalTrials.gov* (NCT03960034). The prospective cohort of lung cancer patients was recruited at the Instituto do Cancer do Estado de São Paulo (ICESP; Sao Paulo, Brazil) between April 2017 and September 2020. Written informed consent was obtained from all participants. All patients presented all the following inclusion criteria: (1) histologically proven diagnosis of stage IV NSCLC; (2) Eastern Cooperative Oncology Group Performance Status between 0 and 2; (3) no previous NSCLC treatment; (4) current or previous smoking habits; (5) normal renal and hepatic functions; (6) able to perform physical tests, according to the investigatorś judgement; and (7) able to read, understand and sign the consent form. The exclusion criteria included (1) any previous systemic treatment for metastatic disease and (2) diagnosis of specific tumor driver mutations, including epidermal growth factor receptor (EGFR) mutations and anaplastic lymphoma kinase (ALK) translocations. A total of 1079 patients were admitted and screened in the hospital, and a final cohort of 55 NSCLC patients was included in the data analysis (**Figure 1**). Additionally, seven age-matched control subjects were recruited. All control subjects were smokers and did not present a previous diagnosis of NSCLC or any other cancer.

Clinical characteristics and body composition were assessed on the day of inclusion, including age, sex, body mass index (BMI), histological type of tumor, ECOG PS, and smoking status. Cancer cachexia was determined based on loss of body mass in the previous six months ^30^, and quality of life (QoL) was assessed using the EORTC-QLQ-C30 ^31^, EORTC-QLQ-CAX24^32^, and SF36 ^33^ questionnaires. Physical performance was determined with a series of functional tests, including the timed up and go test ^34^, the sit-to-stand test ^35^, and the six-minute walking test (6MWT) ^36^. A subcohort of 23 patients was also subjected to an ergospirometry test ^37^. Skeletal muscle and adipose tissue area were analyzed using computed tomography (CT) from the third lumbar vertebra previously collected while determining the NSCLC stage ^38^. Blood samples were collected, and circulating hematocrit levels, hemoglobin, lymphocytes, and neutrophils were determined. Finally, the overall survival rate was determined, and additional blood samples were collected from a subcohort of 18 patients for further studies in cell culture.

### Quality of life (QoL)

QoL was assessed using the EORTC-QLQ-C30 ^31^, EORTC-QLQ-CAX24 ^32^, and SF36 ^33^ questionnaires, with validated versions for the Brazilian population. The EORTC QLQ-C30 was developed to evaluate the quality of life in cancer patients and contains five functional measures (physical, role, emotional, social, and cognitive), eight symptom measures (fatigue, pain, nausea/vomiting, appetite loss, constipation, diarrhea, insomnia, and dyspnea), global health/QoL, and financial impact. Most items use a 4-item scale, with raw scores transformed to 0-100 and higher scores representing better functioning/QoL and symptom burden ^31^. The EORTC QLQ-CAX24 is a cachexia-specific questionnaire comprising five multi-item scales (food aversion, eating and weight-loss worry, eating difficulties, loss of control, and physical decline) and four single items. All items use a 4-item scale with raw scores being transformed to a 0-100 scale, and higher scores indicate more complications, except for item 54 ^32^. The SF36 was developed to measure general self-reported physical and mental health, comprising 36 questions and eight domains: bodily pain, vitality, role-emotional, role-physical, general health, social function, and mental health ^33^.

### Body composition

In addition to body mass and BMI analysis, CT scans previously collected while determining the NSCLC stage were used to determine skeletal muscle and adipose tissue area. The cross-sectional area of the abdominal CT scan at the level of the third lumbar vertebra (L3) was used to measure the skeletal muscle index (SMI), psoas index, visceral adipose tissue (VAT), subcutaneous adipose tissue (SAT), and intramuscular adipose tissue (IMAT) ^38^. Image analysis software (sliceOmatic V5.0; TomoVision, Magog, QC, Canada) was used to measure specific tissue demarcation by the Hounsfield unit (HU). The skeletal muscle index comprised the psoas, erector spinae, quadratus lumborum, transversus abdominis, internal oblique, external oblique, and rectus abdominis. The L3 skeletal muscle and adipose area were normalized by height squared (cm^2^). Cancer cachexia was diagnosed according to Fearon et al., 2011.

### Physical Performance

Physical performance was determined with the timed up and go test ^34^, sit-to-stand test ^35^, six-minute walking test (6MWT) ^36^, and ergospirometry test ^37^. The timed up-and-go and sit-to-stand tests were developed to measure mobility, leg strength, and endurance in the elderly and other populations ^34,39,40^. The 6MWT measured endurance capacity and was performed according to the American Thoracic Society Statement (2002). An ergospirometry test ^37^ was performed on an Ergoline Spirit 150-cycle ergometer (Bitz, Germany). A ramp protocol and work rate increments of 5 to 10 W every minute until exhaustion or when a maximal respiratory exchange ratio of >1.10 was achieved despite verbal encouragement. Breath-by-breath gas exchange analysis was used with a VMax 229 computerized system (SensorMedics). Heart rate and blood pressure were also continuously recorded during the ergospirometry test.

### Cell culture

Cell culture experiments were performed in differentiated human myotubes, except for DR4 luciferase activity, which was performed in C2C12 myoblasts. Primary human skeletal myoblasts (Thermo Fisher Scientific; A11440) were cultured with SkGM™-2 Skeletal Muscle Cell Growth Medium-2 Bullet Kit (Lonza, CC-3245) supplemented with 10% fetal bovine serum (FBS) and 1% pen/strep. Myoblasts were differentiated into myotubes over four days by replacing media with Dulbecco’s modified Eagle’s medium (DMEM; Gibco) supplemented with 2% horse serum and 1% pen/strep. After five days of differentiation, myotubes were maintained in SkGM™-2 Skeletal Muscle Cell Growth Medium-2 Bullet Kit (Lonza, CC-3245) supplemented with 10% FBS or patient plasma and 1% pen/strep. C2C12 cells were cultured in DMEM supplemented with 10% FBS and 1% pen/strep. To challenge myotubes/myoblasts with human plasma samples, FBS was replaced with 10% (initial experiments) or 1.5% human plasma. Endpoint readouts were performed after 16 hours of treatment, including oxygen consumption rate (OCR), extracellular acidification rate (ECAR), cell proliferation, protein content, f-actin expression, and DR4 luciferase activity.

OCR and ECAR were measured with extracellular flux analysis (XF96, Agilent Seahorse, MA USA) in sodium bicarbonate-free DMEM supplemented with 31.7 mM NaCl, 10 mM glucose, and 2 mM glutamax (pH 7.4 adjusted using NaOH) as previously described (C. R. R. Alves et al., 2020; Townsend et al., 2013).. Cell viability was measured with the MTS cell proliferation assay (CellTiter 96 AQueous One Solution Cell Proliferation Assay MTS, Promega). The MTS tetrazolium compound (20 _μ_L) was added to each well, and plates were incubated for an additional 3 h at 37 °C before reading absorbance at 490 nm. Total protein content was measured with a BCA assay (Thermo Fisher Scientific, 23225). Immunofluorescence was performed as previously described ^13,16,42^ to stain F-actin and nuclei in myotubes using Alexa Fluor 488 phalloidin (Thermo Fisher Scientific; A12379) and DAPI (Thermo Fisher Scientific; 62248), respectively. F-actin expression and nuclei count were quantified using IMAGE J ^43^. The Dual-Luciferase Reporter Assay System (Promega) was utilized to measure DR4 luciferase activity (firefly), and normalization in each well was performed using Renilla luciferase internal control as previously described ^13^.

### Liquid chromatography–mass spectrometry (LC-MS) metabolic profiling

Plasma samples from a subcohort of 17 NSCLC patients and seven healthy control subjects were used for metabolic profiling using LC-MS methods as previously described ^44^.

### Statistical analysis

Each outcome was divided into terciles (high, intermediate, and low). Kaplan-Meier curves and log-rank tests were used to analyze the probability of survival among terciles. Two-way analysis of variance (ANOVA) was employed when comparing four experimental groups, followed by Fisher’s least significance difference test for multiple comparisons. An unpaired Student’s t-test was used when comparing two experimental groups. A false discovery rate (FDR) of 10% was set in the metabolomic analysis. A Pearson correlation coefficient (r) was employed to verify the correlation between the metabolites in patient plasma. Pathway enrichment was performed using Metaboanalyst version 5.0 (https://www.metaboanalyst.ca/). Statistical analyses were conducted using Graph Pad Prism 8 (Graph Pad Software Inc., USA), and the statistical significance was set at p < 0.05.

## Data Availability

All data produced in the present work are contained in the manuscript

## Acknowledgment

We are grateful to all participants who participated in this study. We thank Fabiana Hirata and Rodrigo Polizio for technical assistance with clinical tests. This work was supported by a grant from Fundação de Amparo à Pesquisa do Estado de São Paulo (FAPESP; 15/22814-5). WN and CRRA received fellowships from FAPESP (16/20187-6, 19/17009-7; 14/03016-8; 16/01478-0). PCB was supported by a grant from Conselho Nacional de Pesquisa e Desenvolvimento (CNPq; 308071/2021-2).

## Authors’ contributions

WN, CRRA, PCB, KS, and GCJ conceived and designed the study. WN, GS, and JJNA collected clinical data. WN and CA performed cell culture experiments. AD, KP, CD, LB and CBC performed metabolomic analysis. WN and CRRA performed the data analysis and drafted the manuscript. All authors interpreted the data, participated in the manuscript review, and approved the final manuscript.

---

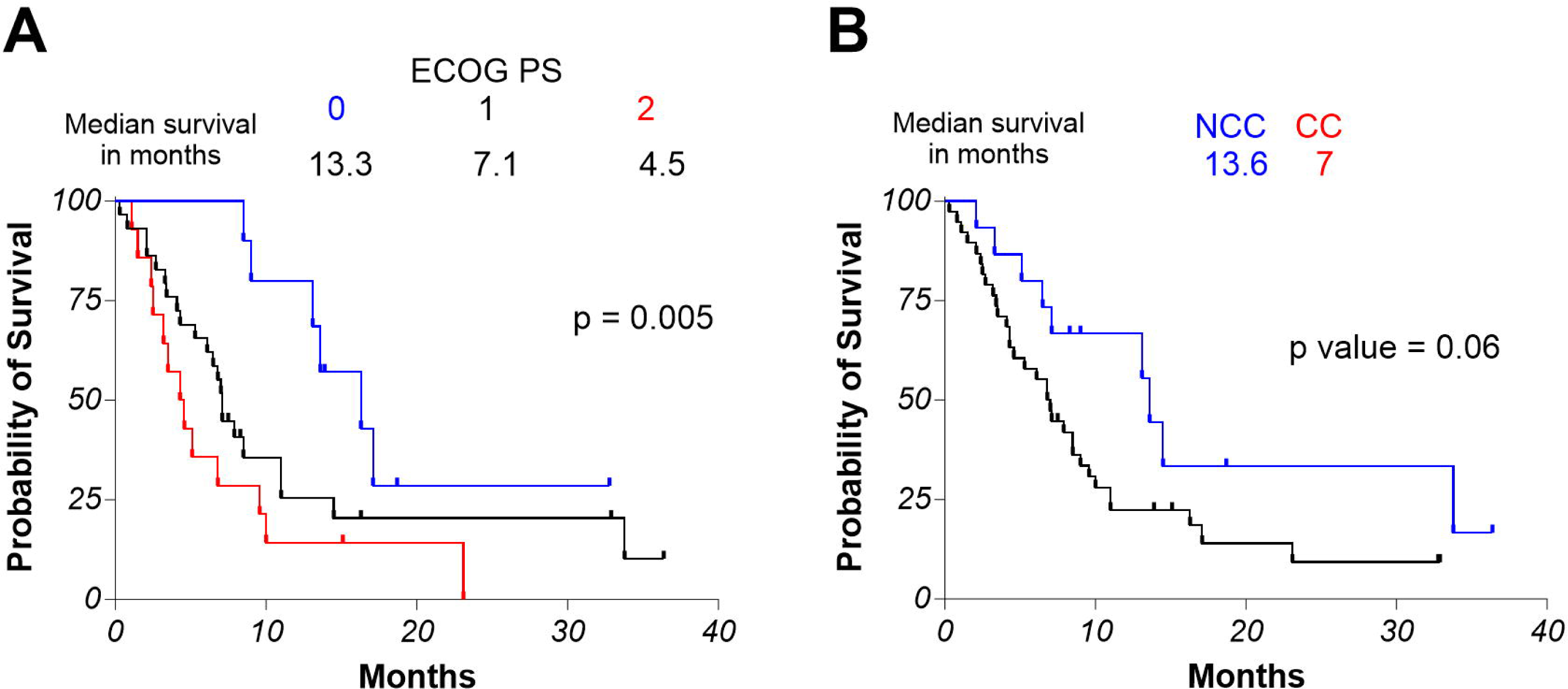

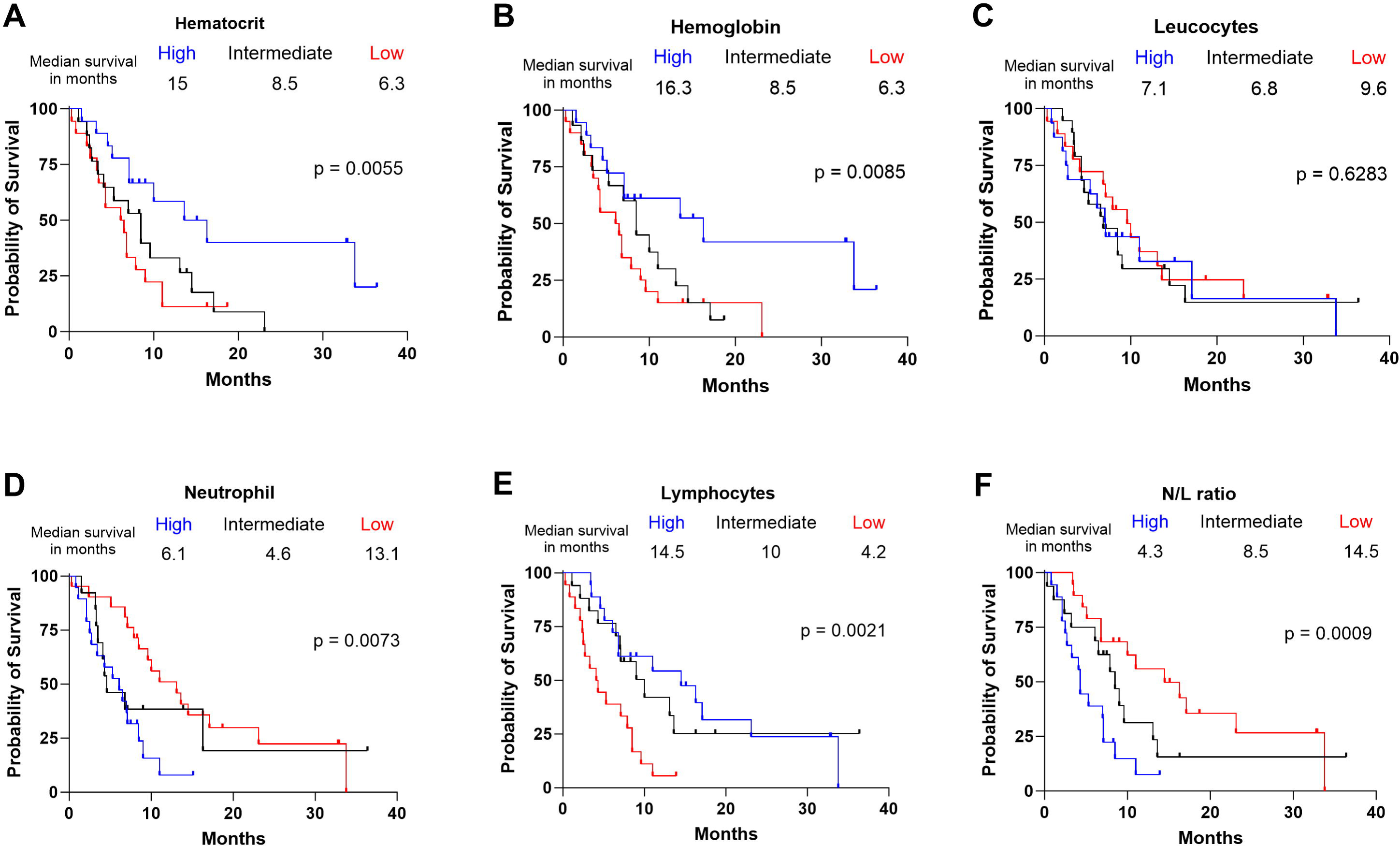

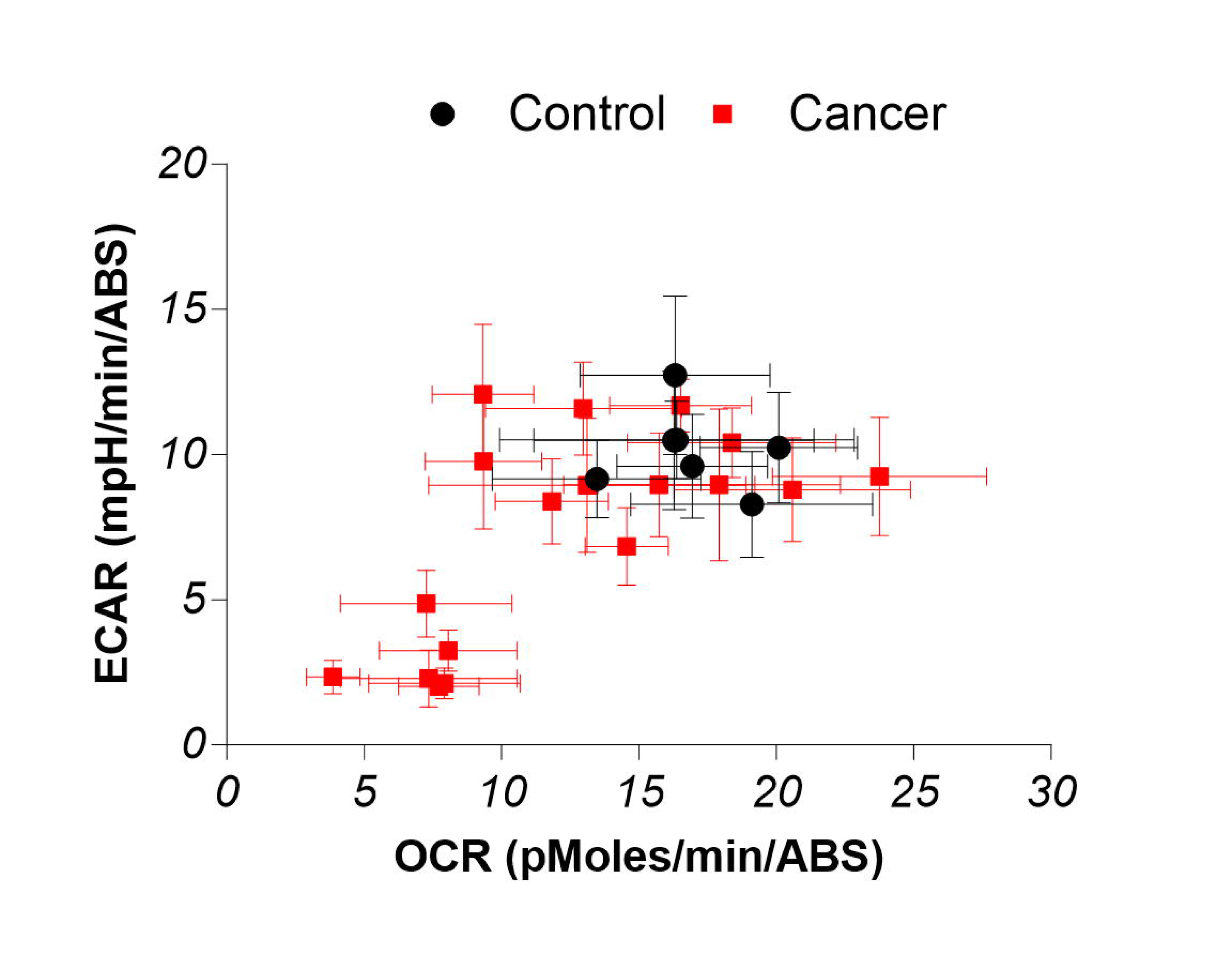

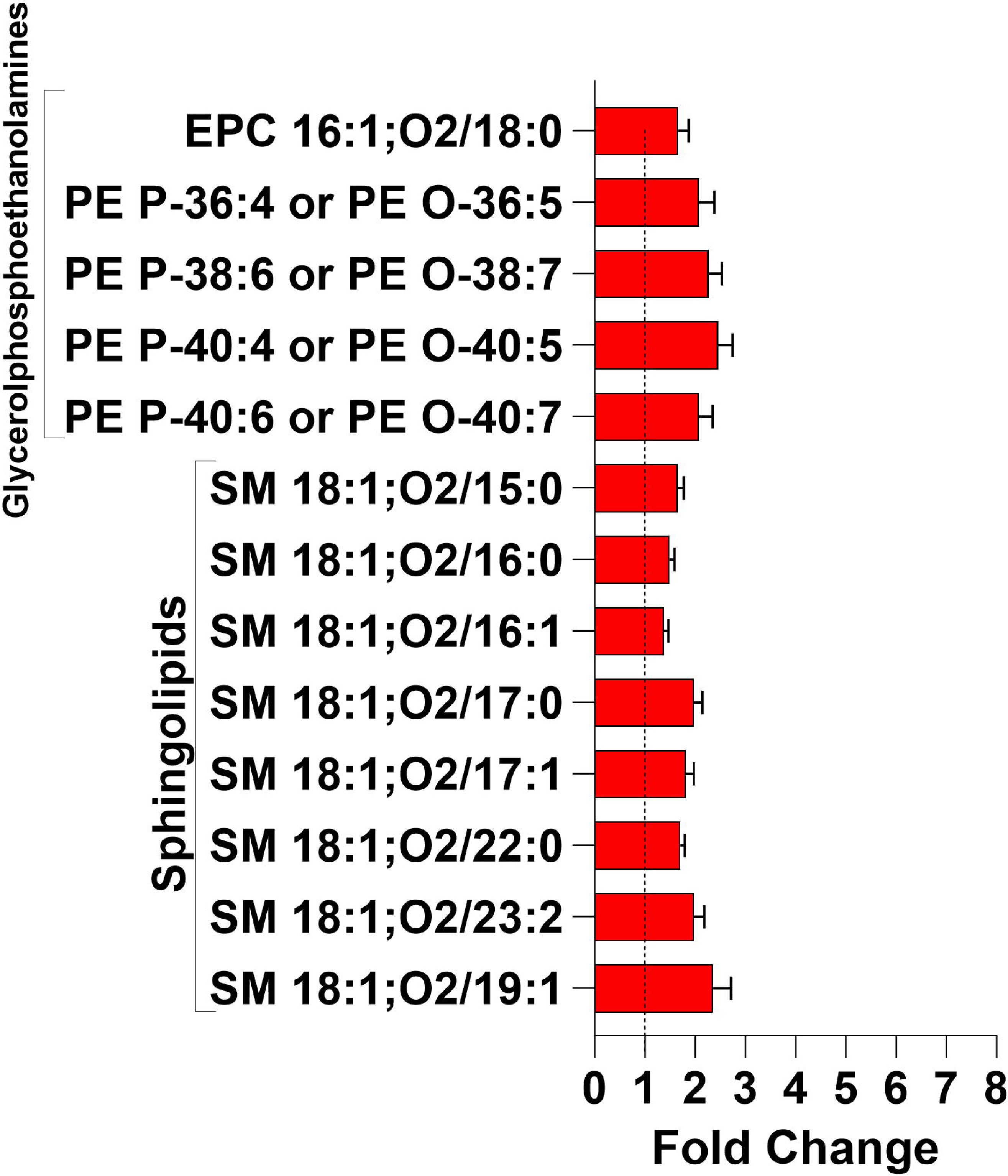

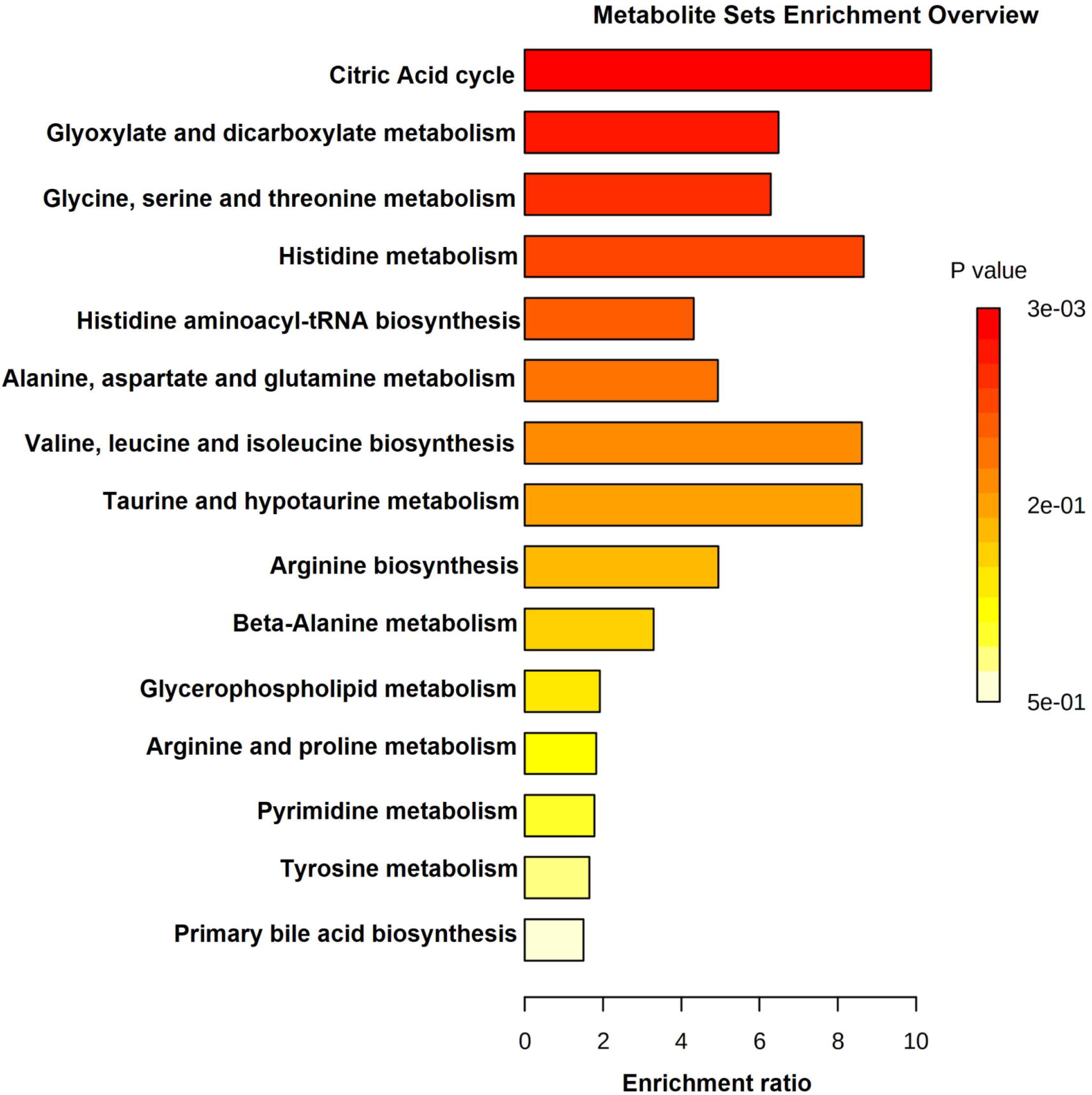

