## Supplementary Note and supplementary tables for "Circulating metabolites and physical performance are predictors of overall survival in metastatic lung cancer patients"

**Supplementary Materials**

**Supplementary Note**

Since physical capacity is typically associated with blood cell counts, we examined whether blood cell count would also be associated with the mortality rate in NSCLC patients. We analyzed the circulating levels of hematocrit, hemoglobin, leucocytes, neutrophils, and lymphocytes and divided patients into terciles based on the quantification of each one of these markers. From this list, hematocrit, hemoglobin, neutrophil, lymphocyte levels, and neutrophil/lymphocyte ratio were statistically associated with the mortality rate (**Figure S2A-F**). Additional measurements analyzed in this study that were not significantly associated with the mortality rate are displayed in **Table S1**.

In addition, we tested potential direct correlations between physical tests, quality of life, and blood cells (**Table S2**), independent of the mortality rate analysis. As expected, we found associations between different physical tests, as well as between physical tests and blood cell counts (**Table S2**). These data indicate that blood cell counts may be associated with worse overall survival rates in NSCLC patients, similar to our observation of physical performance.

**Table S1.** Associations between complementary variables, including quality of life questionaries and probability of survival in non-small cell lung cancer (NSCLC) patients.

| **Measurement** | **Median of survival in months** | | | **p-value*** |
| --- | --- | --- | --- | --- |
|  | High | Intermediate | Low |  |
| Visceral adipose tissue | 13.1 | 7 | 11 | 0.5251 |
| Subcutaneous adipose tissue | 10 | 6.9 | 8.5 | 0.4714 |
| Platelets | 8.5 | 11 | 5.3 | 0.9503 |
| Creatinine | 13.1 | 6.9 | 7.5 | 0.5174 |
| Global quality of life | 9 | 8.2 | 6.8 | 0.1243 |
| Global Health status | 10 | 6.1 | 7.5 | 0.9993 |
| Appetite worries | 7 | 7.5 | 9 | 0.4671 |
| Food aversion | 7 | 6.1 | 9.6 | 0.0680 |
| Vitality | 6.8 | 8.9 | 11 | 0.3315 |
| MVPA | 14.5 | 6.3 | 6.8 | 0.1993 |
| Steps per day | 12.3 | 7.6 | 5.7 | 0.2356 |
| *p-value is related to log-rank test to the survival curves | | | | |

**Table S2.** Correlation between physical tests, quality of life, and blood cell count.

| **Variable** | **95% confidence interval** | **R** | **p-value** |
| --- | --- | --- | --- |
| VO2peak vs 6MWT | 0.2881 to 0.8639 | 0.665 | 0.002 |
| VO2peak vs Sit-stand test | -0.7761 to 0.1112 | -0.518 | 0.016 |
| Sit-stand test vs Handgrip | -0.5336 to 0.0325 | -0.274 | 0.079 |
| Sit-stand test vs TUG | 0.6229 to 0.8606 | 0.766 | <0.000 |
| TUG vs 6MWT | -0.8936 to -0.6829 | -0.813 | <0.000 |
| TUG vs Handgrip | -0.6372 to -0.1329 | -0.416 | 0.005 |
| Sit-stand test vs 6MWT | -0.7018 to -0.2599 | -0.514 | 0.000 |
| 6MWT vs Handgrip | 0.04327 to 0.6204 | 0.366 | 0.027 |
| 6MWT vs Fatigue | -0.4860 to 0.1218 | -0.201 | 0.219 |
| TUG vs Fatigue | -0.0414 to 0.5052 | 0.251 | 0.092 |
| Sit-stand test vs Fatigue | 0.2122 to 0.6753 | 0.476 | 0.000 |
| Sit-stand test vs Physical decline | 0.3070 to 0.7269 | 0.550 | <0.000 |
| 6MWT vs Physical decline | -0.4276 to 0.1939 | -0.129 | 0.432 |
| TUG vs Physical decline | -0.0161 to 0.5238 | 0.275 | 0.064 |
| 6MWTvsPhysical role | -0.1999 to 0.4545 | 0.142 | 0.413 |
| TUG vs Physical role | -0.4771 to 0.1246 | -0.194 | 0.229 |
| Sit to stand vs Physical role | -0.5138 to 0.0764 | -0.240 | 0.135 |
| 6MWT vs Physical health summary | -0.1901 to 0.4541 | 0.147 | 0.390 |
| TUG vs Physical health summary | -0.5027 to 0.0827 | -0.230 | 0.147 |
| Sit to stand vs Physical health summary | -0.5839 to -0.0324 | -0.336 | 0.032 |
| 6MWT vs N/L ratio | -0.4343 to 0.1388 | -0.161 | 0.289 |
| TUG vs N/L ratio | -0.1096 to 0.4219 | 0.168 | 0.233 |
| Sit to stand vs N/L ratio | -0.0110 to 0.5041 | 0.265 | 0.059 |
| Sit to stand vs Hemoglobin | -0.5578 to -0.0637 | -0.333 | 0.016 |
| Tug vs Hemoglobin | -0.4429 to 0.0839 | -0.193 | 0.169 |
| 6MWT vs Hemoglobin | -0.1155 to 0.4533 | 0.184 | 0.226 |
| 6MWT vs Hematocrit | -0.1403 to 0.4331 | 0.159 | 0.294 |
| TUG vs Hematocrit | -0.4083 to 0.1257 | -0.152 | 0.281 |
| Sit to stand vs Hematocrit | -0.5578 to -0.0637 | -0.333 | 0.016 |
| Sit to stand vs Leucocytes | -0.0495 to 0.4748 | 0.229 | 0.106 |
| TUG vs Leucocytes | -0.1388 to 0.3971 | 0.139 | 0.325 |
| 6MWT vs Leucocytes | -0.4020 to 0.1769 | -0.123 | 0.421 |
| 6MWT vs Lymphocytes | -0.1083 to 0.4591 | 0.191 | 0.208 |
| Sit to stand vs Lymphocytes | -0.4100 to 0.1294 | -0.151 | 0.288 |
| Sit to stand vs Neutrophils | 0.0048 to 0.5293 | 0.288 | 0.046 |
| TUG vs Lymphocytes | -0.4501 to 0.0750 | -0.202 | 0.151 |
| TUG vs Neutrophils | -0.0528 to 0.4816 | 0.231 | 0.109 |
| 6MWT vs Neutrophils | -0.4904 to 0.0908 | -0.219 | 0.163 |

**Table S3**. Demographic characteristics of a subset of NSCLC patients and healthy control subjects with plasma samples available for further in vitro studies with myotubes.

| **Characteristics** | **Control** | **Cancer** |
| --- | --- | --- |
| **N** | 7 | 18 |
| **Age** (mean ± SD) | 60 (4.7) | 63 (8.1) |
| **Sex** (N) |  |  |
| Male | 7 | 16 |
| Female | 0 | 2 |
| **Weight** (mean ± SD) | 61.4 (57) | 65.1 (16.2) |
| **BMI** (mean ± SD) | 22.2 (3.4) | 22.3 (4.7) |

**Figure 1.** Flowchart detailing subject selection. NSCLC: non-small cell lung cancer.

**Figure S1: Overall survival analysis**. (**A**) ECOG-PS status (0, 1, and 2) was plotted in a Kaplan-Meier survival curve and compared using a log-rank test. (**B**) Kaplan Meier curve comparing cachectic and non-cachectic patients.

**Figure S2. Associations between blood cells and probability of survival in non-small cell lung cancer (NSCLC) patients.** Each measurement was divided into terciles (high, intermediate, and low), and terciles' probability of survival was compared. Blood cells analysis included (**A**) Hematocrit percentage**,** (**B**) Hemoglobin, (**C**) Leucocytes, (**D**) Neutrophil, (**E**) Lymphocytes, (**F**) Neutrophil to lymphocytes ratio. Comparisons were performed using the log-rank test and presented as Kaplan-Meier curves. n = 55.

**Figure S3. Metabolic changes in primary human myotubes treated with plasma from NSCLC patients**. Differentiated primary human myotubes were incubated for 16 hours with media containing 10% plasma collected from each subject. Oxygen consumption rate (OCR; x-axis) and extracellular acidification rate (ECAR; y-axis) were normalized by protein content and plotted.

**Figure S4. Expression of sphingolipids and glycerolphosphoethanolamines**. Fold change between NSCLC patients with low endurance capacity and control subjects.

**Figure S5.** KEGG pathway analysis using all differentially expressed metabolites identified in main Figure 5.
